# Large Language Models Generate Stigmatizing Language During Reasoning Over Real-World Clinical Data

**DOI:** 10.64898/2026.08.12.26360210

**Authors:** Yutong Yang, Bowen Gu, David B. Hathaway, Richard Wyss, Laura Marengo, Jason B. Gibbons, Stanley Lyndon, Jiageng Wu, Qingyu Chen, Nan Liu, Philip S Wang, Leo Anthony Celi, David W. Bates, Kueiyu Joshua Lin, Li Zhou, Jie Yang

## Abstract

Stigmatizing language in clinical documentation, which conveys negative stereotypes, attitudes, or judgments toward patients, is a recognized source of documentation bias and is associated with poorer care and adverse health outcomes. Although prior stigma-related research has explored on clinician-written EHR notes, the increasing use of large language model (LLM)-generated documentation in clinical workflows raises new concerns about its potential to produce or amplify bias and affect patient safety. In this study, we conducted a large-scale assessment of stigmatizing language in LLM-generated reasoning text on 35 real-world clinical tasks across 107 LLMs. We applied a psychiatrist-validated, natural language processing (NLP) system to detect stigma terms in LLM reasoning text and quantified stigma rates of LLM-generated reasoning texts across 3,745 model-task pairs. Results showed that stigma rates ranged from 0% to 33.33%, with 84.06% of pairs containing stigma terms. Reasoning models showed higher stigma rates than non-reasoning models (2.35% vs. 1.70%; p < 0.0001), whereas medical models did not show significantly lower stigma rates than general-purpose models (1.80% vs. 2.00%; p = 0.26). Stigma rates of LLM outputs correlated negatively with task accuracy (r = -0.283; p < 0.001) and positively with input clinical-text stigma (r = 0.569; p < 0.001), with 19.76% of LLM-task pairs *amplifying* stigma in the original input notes. The effectiveness of prompt engineering as a destigmatizing approach varied across models, with stigma rates reduced by up to 91.91% without compromising model performance. This study shows that stigmatizing language generation is common but modifiable in LLM-generated reasoning traces, highlighting the need for direct stigma-related safety evaluation and robust destigmatization strategies before clinical deployment.

## Introduction

Stigmatizing language, which refers to language that communicates unintended meanings that can perpetuate socially constructed power dynamics and result in bias^1^, has been commonly identified in clinical documentation, particularly electronic health records (EHRs)^2–4^ and has been associated with poorer quality of patient care and higher rates of adverse health outcomes^5,6^. In particular, it has been found to impact care through multiple pathways, including reduced quality of care, increased medical errors, and weakened patient–provider relationships^5,7^. Clinicians exposed to stigmatizing chart notes demonstrate more negative attitudes toward patients and prescribe pain medications less aggressively than those reading neutral language notes^5^. About 10% of patients who review their clinical notes report feeling judged or offended by their physicians’ language^7^. Moreover, stigmatizing language in EHR notes is distributed unevenly across patient populations, with particular concentrations in racial and ethnic minoritized populations and across gender lines. For example, black patients and women receive negative framing more frequently in medical records^8^. This disparity extends across clinical specialties and note types and drives inequity in clinical decision-making^3,4^.

Large language models (LLMs) have been increasingly integrated in healthcare and have been adopted in clinical workflows for note drafting, summarization, phenotyping, communications, and documentation support^9–13^. A 2026 American Medical Association survey shows that 30% of physicians incorporate AI use in the creation of discharge instructions, care plans, and/or progress notes, which is a 50% increase in the last two years^14^. Although this integration may reduce documentation burden, directly incorporating LLM-generated text into clinical records could amplify bias and safety risks, possibly through the generation of stigmatizing language^15–17^. If LLM-generated documentation contains stigmatizing language, it could therefore contribute to clinical harm and further exacerbate existing disparities among disadvantaged patient groups. This concern is further heightened by the rise of LLMs with reasoning ability^18–20^, whose outputs often include intermediate explanatory text before final answers and are therefore substantially longer and may harbor stigmatizing language that is less visible to end users yet still embedded in the generation pipeline. Also, the appearance of stigmatizing language in generated reasoning traces may adversely affect the models’ clinical decisions. Identifying and addressing stigmatizing language across the full spectrum of LLM outputs, including their reasoning traces generated over real-world clinical data, is therefore essential for the safe and equitable deployment of these models in clinical settings.

Existing studies have characterized stigmatizing language and related racial disparities in electronic health records written by physicians, nurses, and other clinicians^3,8,21^. Others have investigated the prevalence of stigmatizing language across health systems^4^, explored how biased training corpora can propagate bias in healthcare AI systems^16,22^, highlighted challenges in under-detecting stigmatizing language^23^, examined LLMs’ stigmatizing behavior in contextual judgements^24^, and developed machine learning approaches with ClinicalBERT^25^ for identifying stigmatizing and preferred language in clinical notes^26^. However, to our knowledge, no study has examined stigmatizing language in *LLM-generated clinical text*. This reveals an urgent evidence gap as LLMs are increasingly integrated into clinical documentation workflows and clinical decision support.

In this work, we systematically evaluated whether LLMs generate stigmatizing language during their reasoning traces over processing large-scale real-world clinical data. We examined model reasoning text from 107 LLMs on 35 real-world clinical tasks under the Chain-of-Thought (CoT) inference setting^18^. We then applied a psychiatrist-validated NLP system to identify stigma terms throughout each model’s reasoning text^4^. We compared the stigma rates, defined as the number of LLM-generated reasoning texts with at least one stigma term over all LLM generations, and stigma term distribution across different models and task characteristics, and quantified correlations between the stigma rates of LLMs’ reasoning text and corresponding input text. Then, we examined the change of stigma rates under tasks with different accuracies. Finally, we explored whether the stigma rates can be reduced and whether the reduction comes at a cost to the model performance. This evaluation addresses the gap in understanding whether LLMs generate stigmatizing language on real-world clinical data and how it can be mitigated, with implications for destigmatizing approaches, deployment safeguards, health equity, and patient safety.

## Results

Through the evaluation against two independent psychiatrists’ review (D.B.H and L.M.) of a stratified random sample of 200 sentences (details in **Appendix 6**) with high inter-rater agreement (Cohen’s κ = 0.96), the stigma-detection NLP system^2–4^ achieved a positive predictive value (PPV) of 94% and a negative predictive value (NPV) of 100%, showing its credibility in identifying stigma terms.

### Overall Distributions of Stigma Text

The overall distribution of all models’ reasoning traces stigma rate across all real-world English clinical tasks is shown in **Figure 1**. While most tasks showed low stigma rates across all models, tasks sourced from the MIMIC dataset^27^, a large and open-source de-identified clinical database, resulted in higher stigma rates. On the model side, the medical model Baichuan-M2-32B^28^ showed both the highest single-task stigma rate on the MIMIC-IV CDM task^29^ (33.33%) and the highest average stigma rate (4.24%). By comparison, the best-performing proprietary models, gpt-4o-0806 and gemini-2.5-flash, had average stigma rates of 1.67% and 1.64%, respectively. Among the best-performing open-source models, DeepSeek-R1 and gemma-4-31B-it models were associated with average stigma rates of 2.17% and 1.36%, respectively.

**Figure 1.**
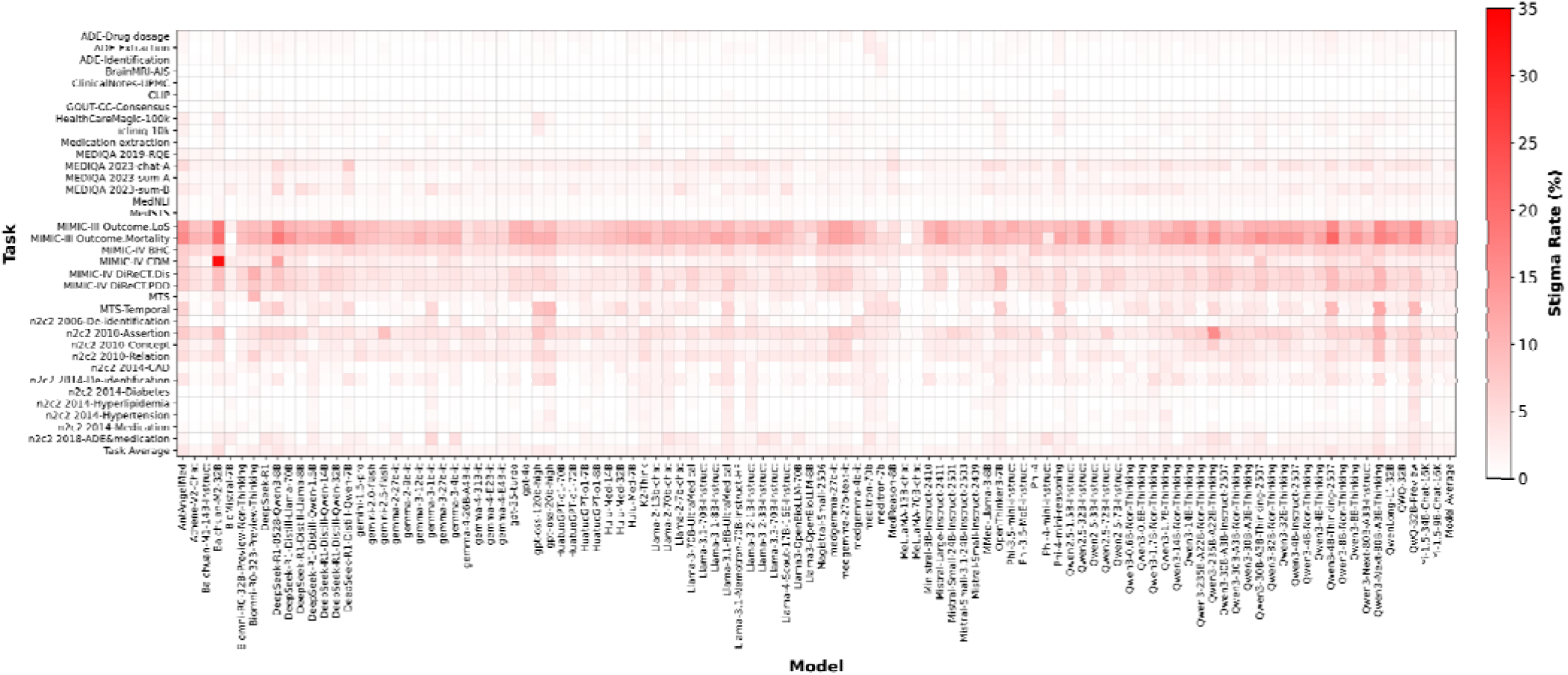
General distribution of stigma rate on models’ reasoning traces across 107 LLMs on 35 tasks. Each cell represents the stigma rate for a specific model–task combination, with red indicating higher stigma rates.

**Figure 2** shows the stigma rate across model–task pairs, clinical applications, task types, and source clinical document types. Stigma rates across all 3,745 model–task pairs (107 LLM models × 35 tasks) ranged from 0.00% to 33.33% (**Figure 2A**). Among all pairs, 3,148 pairs (84.1%) showed positive stigma rates, with an overall mean rate of 2.32% and a median of 1.28%. High (≥5%) stigma rates were observed in 396 pairs (12.58% of positive pairs), while low stigma rates (<1%) occurred in 1,300 airs (41.30% of positive pairs). **T**he mean stigma rates also varied across different clinical applications (**Figure 2B**). Prognosis prediction had the highest mean rate (9.55%), followed by summarization (5.07%). The lowest stigma rate was in negation identification (0.01%). Other clinical applications had intermediate stigma rates (0.53%-2.64%; see **Appendix 4** for the per-task stigma rate).

**Figure 2.**
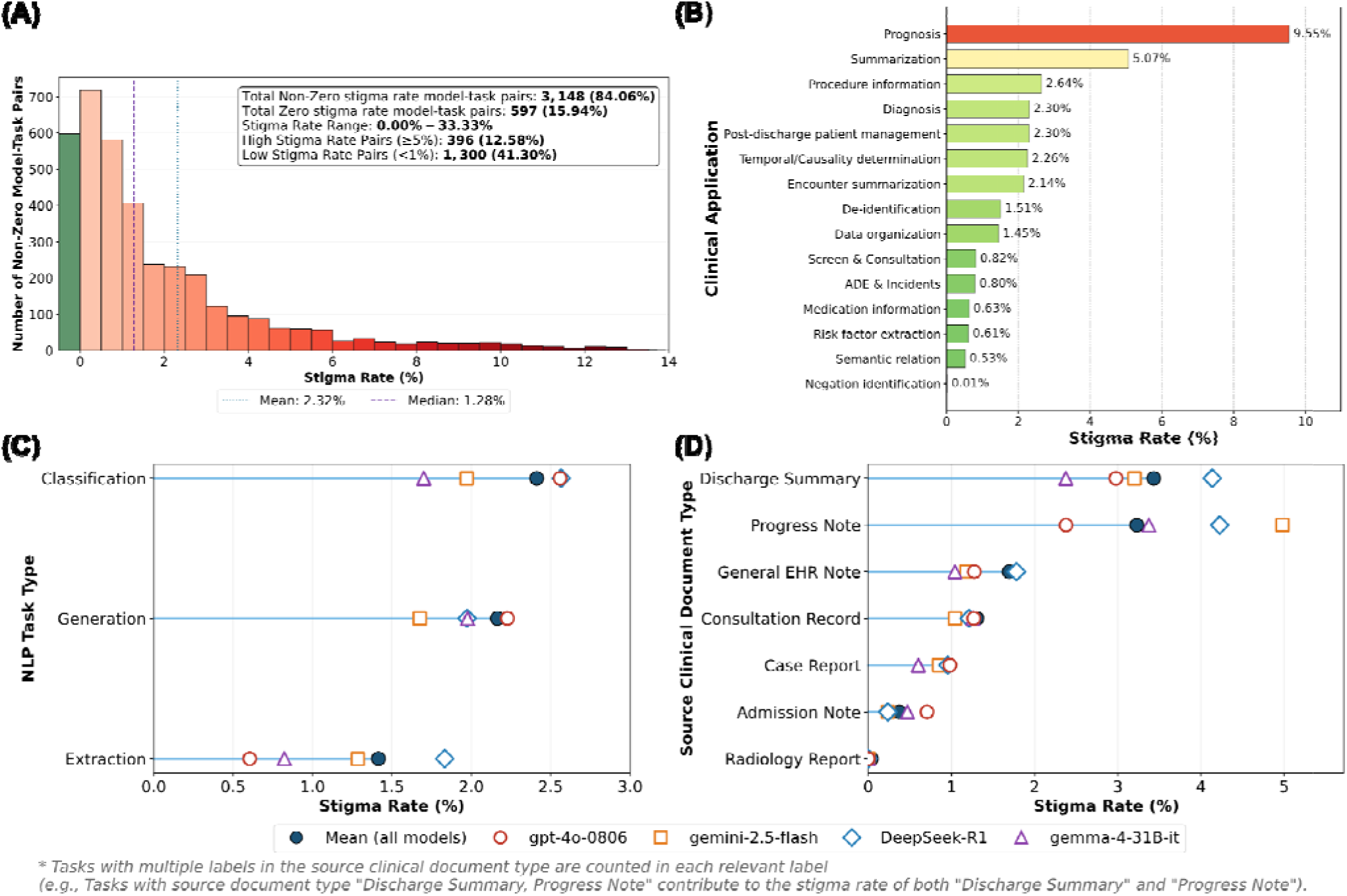
Stigma distribution across model–task pairs, clinical applications, task types, and source clinical document types. (A) Distribution of stigma rates across all model–task pairs. The green bar indicates the zero stigma rate model-task pairs (N = 597). The blue dotted line marks the mean (2.32%), and the purple dashed line indicates the median (1.28%). (B) Mean stigma rate by clinical application. (C) Mean stigma rate of the highlighted models by NLP task types. (D) Mean stigma rate of the highlighted models by source clinical document type. Tasks with multiple labels in the source clinical document type are counted in each relevant label. For detailed definitions of each category under “Clinical Application”, “NLP Task Type”, and “Source Clinical Document Type”, please refer to BRIDGE^34^.

To study model-specific heterogeneity in the use of stigmatizing language, we chose four highlighted models with two advanced proprietary models (gpt-4o-0806^30^, gemini-2.5-flash^31^) and two open-source models (DeepSeek-R1^32^, gemma-4-31B-it^33^). Results showed that all models generated stigmatizing language, and their average stigma rates were strongly affected by task type (**Figure 2C)** and source clinical document type (**Figure 2D**). Classification tasks showed higher stigma rates than generation and extraction tasks (2.41% vs. 2.16% and 1.42%). As for stigma rate across different source clinical document types, LLMs showed the highest mean stigma rate when processing discharge summary notes (3.43%), followed by the progress notes (3.23%), while processing radiology reports led to the lowest stigma rate (0.04%). Deepseek-R1 and gemini-2.5-flash use disproportionately high rates of stigmatizing language in discharge summary and progress notes, respectively. These patterns reflect document structure: narrative-heavy, clinician-authored documents like discharge summaries are more likely to elicit stigma language within models’ reasoning trace, whereas highly structured formats such as radiology reports can reduce stigmatizing language generation.

### Stigma Rates across Model Categories

**Figure 3A** presents differences in model reasoning traces stigma rates across the three model category groups based on Welch’s t-test. Reasoning models (n = 41) had a higher mean stigma rate than non-reasoning models (n = 66) (2.35% vs. 1.70%; p < 0.0001). Open-source models (n = 102) exceeded proprietary models (n = 5) (1.97% versus 1.60%; p < 0.01) in stigma rates. Stigma rates between general models (n = 81) and medical models (n = 26) did not differ significantly (2.00% versus 1.80%; p = 0.26). We note that the statistical power of the comparison between the open-source and proprietary models is limited, given the limited sample size of proprietary models.

**Figure 3.**
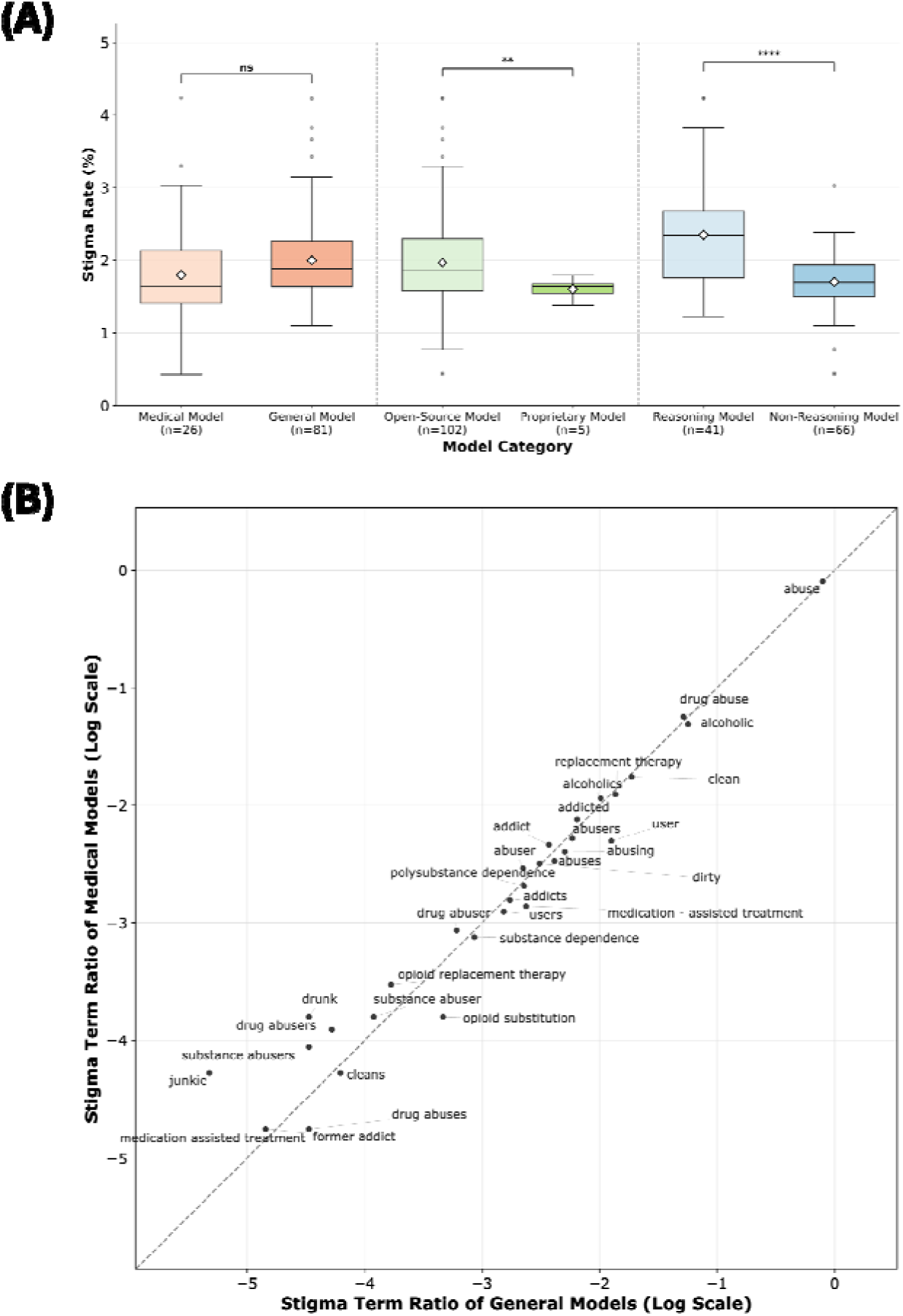
Stigma patterns in model reasoning traces across model categories. (**A**) Box plot of stigma rates by the three model category groups based on Welch’s t-test. “ns” indicates that the stigma rate difference is not statistically significant (p ≥ 0.05). “**” indicates that the stigma rate difference is statistically significant with p < 0.01. “****” indicates that the stigma rate difference is statistically significant with p < 0.0001. The grey dashed lines are the dividers of different model category groups. (B) Stigma term proportion comparison between the general and medical models. Here, “proportion” refers to the ratio of each stigma term across all stigma terms detected, grouped by the model domain (medical vs. general). The log scale is used to narrow the proportion gap between the most and least frequent terms. The grey dashed line indicates an equal proportion. For the log scale, log base 10 is used.

### Difference of Stigma Terms between Medical and General Models

**Figure 3B** shows the distribution of the stigma terms (**Appendix 2**) between the general and medical models. We noticed that for most terms, especially the terms that appear the most frequently (top right corner), the distribution is very close between the two model groups. For example, the term “abuse” has general: medical model proportion = 1: 1.02, “alcoholic” is 1: 0.86, and “drug abuse” is 1: 1.09. For terms with lower frequency, the medical models used “drug abusers” (1: 2.21) and “substance abusers” (1: 2.45) at a higher rate than the general models, whereas the general models used terms such as “user” (2.54: 1) and “opioid substitution” (3.26: 1) more frequently than the medical models.

### Stigma Rates across Model Families

We selected four LLM families to study the relationship between models’ reasoning traces stigma rate and model scale. As shown in **Figure 4A**, the common scaling-law assumption^35^ that larger models achieve better performance does not hold consistently across all tested model families, as the stigma rate does not decrease as the model parameter increases. This indicates that generating stigmatized language is a consistent issue across all model sizes and will not be mitigated by expanding model parameters.

**Figure 4.**
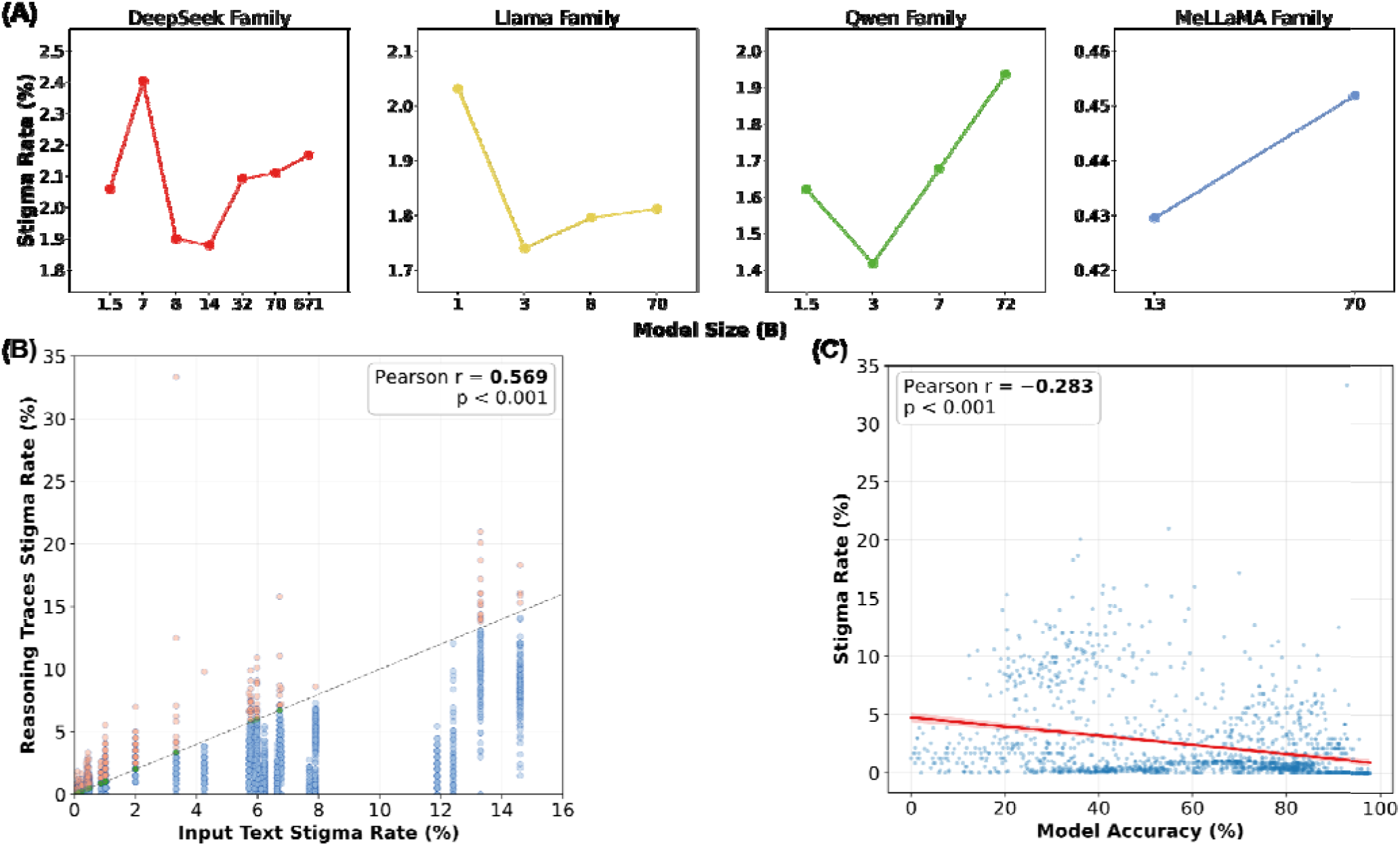
Stigma rates in model reasoning traces and their associations with input text and model performance. **(A)** Reasoning traces stigma rates by model family. The “DeepSeek Family” includes the DeepSeek-R1-Distill-Qwen (1.5B, 7B, 14B, 32B), the DeepSeek-R1-Distill-Llama (8B, 70B), and the DeepSeek-R1 (671B) models. The “Llama Family” includes the Llama-3.1 (8B, 70B) and the Llama-3.2 (1B, 3B) models. The “Qwen Family” includes the Qwen-2.5 (1.5B, 3B, 7B, 72B) models. The “MeLLaMA Family” includes the MeLLaMA (13B, 70B) models. **(B)** Pearson correlation between models’ reasoning traces and input text stigma rates on all 3,745 model–task pairs. Each dot represents one model–task pair. The dashed gray line indicates perfect correlation (y = x). The red dots, green dots, and blue dots indicate the pairs that are above, on, and below the perfect correlation line, respectively. (C) Pearson correlation between reasoning traces stigma rate and model accuracy. Only classification tasks were included (15 out of 35 tasks). Each point on the plot represents one model-task pair (n = 1,605). The red solid line is an ordinary least squares (OLS) linear regression line, with the red shadowed area showing the 95% CI band for the mean regression line.

### Correlation between Reasoning Traces Stigma Rate and Input Text Stigma Rate

We investigated the Pearson correlation between the reasoning traces stigma and the input text stigma on all 3,745 model-task pairs. The result is shown in **Figure 4B**. We found positive correlations (Pearson r = 0.569; p < 0.001) between the two stigma rates. Among all pairs, 61.17% fell below the perfect correlation line (lower reasoning trace stigma rate than input text stigma rate), 19.76% are above the line (higher reasoning traces stigma rate than input text stigma rate), and 19.07% are on the line (the two stigma rates are equal). Notably, we found that LLMs can generate stigmatizing language in their reasoning traces even when the corresponding input text does not contain stigma terms. For example, in the task ClinicalNotes-UPMC (negation identification)^36^ and the task BrainMRI-AIS^37^ (acute ischemic stroke diagnosis), none of the input text had stigma terms. However, 10 of the 107 models had positive stigma rates in their reasoning traces.

### Correlation between Reasoning Trace Stigma Rate and Model Accuracy

**Figure 4C** shows the Pearson correlation between reasoning trace stigma rate and model accuracy on the tasks, with an OLS linear regression line fitted to the data and the red shadowed area showing the 95% CI band. We note that only the 15 classification tasks out of the 35 tasks that use accuracy as the evaluation metric are involved, as it is more straightforward to quantify model performance via accuracy. Overall, we identify a moderately negative but statistically significant linear association between the stigma rate and the accuracy (Pearson r = −0.283; p < 0.001), suggesting that higher model accuracy is partially associated with lower stigma rate in the reasoning traces.

### Destigmatizing the output of LLMs

We explored a simple destigmatizing method by instructing the model to avoid generating the stigma terms listed in **Appendix 2** using prompt engineering (**Appendix 5**). **Figure 5** presents the reasoning traces stigma rate and performance change of the five selected models (Mistral-Large-Instruct-2411^38^, gemma-4-31B-it^33^, Llama-3.3-70B-Instruct^39^, gpt-4o^30^, Qwen3-Next-80B-A3B-Instruct^40^) after the destigmatizing process. We found that all models except Qwen3-Next-80B-A3B-Instruct had their stigma rate decrease significantly (gemma-4-31B-it shows a 91.91% stigma rate reduction, Mistral-Large-Instruct-2411 – 88.82%, gpt-4o – 86.83%, Llama-3.3-70B-Instruct – 76.24%) after inferencing on the prompt that specifies how to avoid stigma term generation (**Figure 5A**). However, Qwen3-Next-80B-A3B-Instruct had a 26.84% stigma rate increase with the destigmatizing prompt. Further analysis indicated that this divergent response was driven by task type, with Qwen3-Next-80B-A3B-Instruct reducing stigma rates in 12 of 15 classification tasks while increasing them in 11 of 15 extraction tasks. **Figure 5B** demonstrates that modifying the prompt with destigmatizing instructions does not affect the performance of most models, as the performance difference is very small for all models except gpt-4o on information extraction tasks (-3.62% in F1-event after destigmatizing). Overall, we found that the destigmatizing process can effectively reduce the stigma rate of the LLMs while retaining performance.

**Figure 5.**
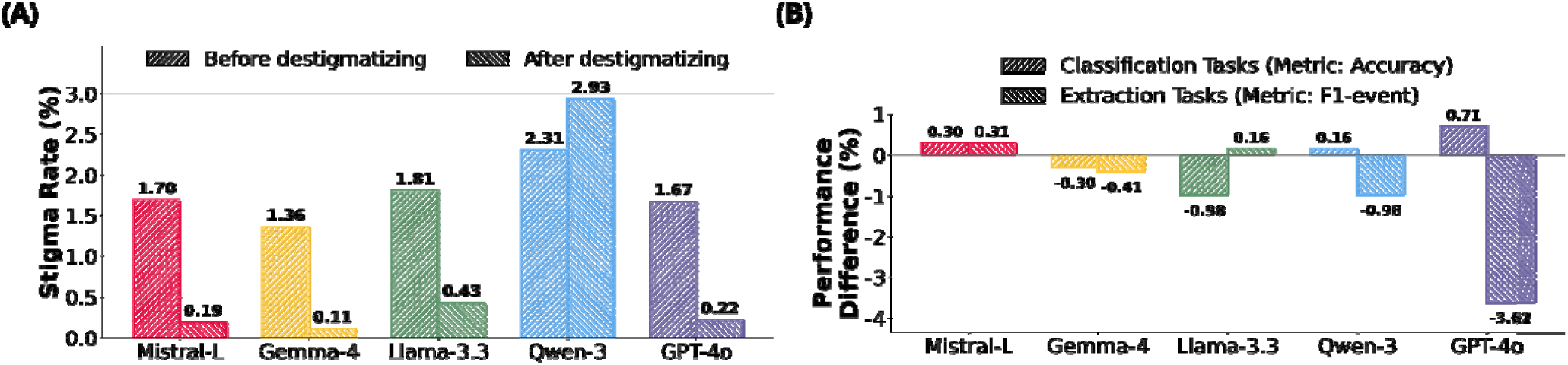
Effects of the destigmatizing step on reasoning-trace stigma and model performance. (A) The change of reasoning traces stigma rate. (B) The change of model performance. The y-axis represent the absolute performance difference. A positive value indicates improved performance after destigmatizing, and vice versa. Abbreviations: Mistral-L - Mistral-Large-Instruct-2411, Gemma-4 - gemma-4-31B-it, Llama-3.3 - Llama-3.3-70B-Instruct, Qwen-3 - Qwen3-Next-80B-A3B-Instruct.

### Frequent Stigma Terms and Language Patterns

We counted the most frequently detected stigmatizing terms across all models’ reasoning traces. We identified 31 distinct stigma terms in total. Stigma terms with the highest appearance frequency were “abuse” (79.79%), followed by “alcoholic” (5.51%), and “drug abuse” (5.31%). Other common terms included “alcoholics”, “addicted”, “abusers”, “abusing”, “abuses”, “addict”, and “dirty”. Most of these strings arise in substance-use, alcohol-use, or medication- and treatment-discussion contexts (**Appendix 2**). Less frequent terms included “polysubstance dependence”, “drug abuser”, “substance abuser”, “drunk”, and “junkie” (each contributing well under 1% of occurrences). This shows that stigmatizing terms in LLM reasoning traces are highly concentrated: the top three and top 10 terms accounting for approximately 90.61% and 97.67% of all occurrences of stigmatizing language, respectively.

## Discussion

In this large-scale evaluation of 107 LLMs across 35 real-world clinical tasks, we found that stigmatizing language was widespread across models and tasks, with heterogeneity in its frequency. Importantly, LLMs did not merely reproduce stigmatizing language present in the source clinical text: they could generate such language when none was detected in the input and amplified stigma relative to the source text in nearly one-fifth of evaluations. We further found that increasing model scale did not consistently reduce stigma across the evaluated model families, and medical models did not exhibit significantly lower stigma than general-purpose models, whereas destigmatizing prompts substantially reduced stigma for several models with little change in task performance. To our knowledge, this is the first large-scale study to systematically evaluate stigmatizing language in LLM-generated reasoning traces across diverse real-world clinical tasks, whereas prior studies have largely focused on final model responses with limited models and clinical settings^24,41^. Our study suggests that stigmatizing language is neither an isolated property of individual models nor an inevitable consequence of processing stigmatized clinical text, but a measurable and potentially modifiable safety concern that should be explicitly evaluated when LLMs are developed and deployed for clinical applications.

Our findings suggest that stigmatizing language generated by LLMs may arise through both the propagation of language present in clinical inputs and model-generated linguistic associations. The positive correlation between stigma rates in source clinical text and model reasoning traces indicates that LLMs can inherit stigmatizing language from the records they process. However, models also generated stigmatizing language when none was detected in the input and amplified stigma relative to the input text. One plausible explanation is that LLMs learn linguistic associations from historical clinical and general-domain corpora in which stigmatizing terminology and biased representations are embedded. Such associations may be reproduced when models interpret or generate clinical text, even in the absence of explicitly stigmatizing input language. If LLM-generated documentation is incorporated into electronic health records without adequate safeguards, historical documentation bias may be reproduced and reintroduced into future training data, creating a feedback loop that could compound risks such as automation bias and potential patient harm^42^.

There were notable differences in stigmatizing language across different model categories. Reasoning models showed higher average stigma rates than non-reasoning models. One possible explanation is that reasoning models typically generate longer and more elaborate reasoning traces, creating more opportunities for stigmatizing terms or associations to appear during generation. Open-source models also showed a modestly higher average stigma rate than proprietary models, although the imbalance in group sizes (102 open-source models vs 5 proprietary models) limits interpretation of this comparison. More notably, medical models did not exhibit significantly lower stigma rates than general-purpose models. This null finding is important because medical domain-specific pretraining or fine-tuning is intended to improve models’ suitability for clinical use, yet our results provide no evidence that medical specialization alone reduces stigmatizing language. Notably, Baichuan-M2-32B is a medical model, while it had the highest average stigma rate among the evaluated models, illustrating that greater clinical specialization does not necessarily translate into safer or more equitable language generation. Therefore, models intended for clinical use should undergo direct evaluation of stigmatizing language and other equity-related harms in the specific tasks and clinical contexts in which they will be deployed.

Our destigmatizing prompt-engineering experiments demonstrate that stigmatizing language is modifiable, with large reductions in stigma rates across several models and minimal changes in task performance. However, the increased stigma rate observed for Qwen3-Next-80B-A3B-Instruct after applying the destigmatizing prompt indicates that prompt engineering should not be regarded as a universal safeguard. More durable mitigation may therefore require model-level interventions, including supervised fine-tuning, preference-based alignment, incorporation of external knowledge to guide clinical reasoning^43^, and additional safeguard training^44^ using examples of clinically appropriate and non-stigmatizing language. Such approaches should aim not simply to suppress individual stigma terms, but to preserve clinically relevant information while reducing judgmental, reductive, or unnecessarily stigmatizing framing. Future work should also examine whether stigmatizing language is associated with changes in model confidence or uncertainty^45^, which may provide additional signals for identifying unsafe or unreliable model behavior. Because the effectiveness of destigmatizing interventions may vary by model and task, mitigation strategies should be validated across the specific clinical applications and document types in which models are intended to be used.

This study has several limitations. First, our evaluation was limited to English-language clinical datasets because the stigma detection system currently supports only English text, which may limit the generalizability of our findings to other languages and cultural contexts. Second, although the rule-based stigma detection system demonstrated high agreement with psychiatrist review, it may miss stigmatizing expressions that are not captured by the predefined terminology or that depend strongly on context. More context-aware approaches, including LLM-based detectors, in-context learning, and domain-adapted classifiers, may help address this limitation^23,26,46^. Third, the reasoning traces evaluated in this study were generated under a Chain-of-Thought prompting setting and may not represent models’ latent internal reasoning processes. Nevertheless, these traces represent intermediate explanatory text that may be exposed to users or incorporated into downstream clinical workflows. Fourth, our mitigation experiments were limited to prompt engineering in a selected set of models and did not systematically evaluate more advanced approaches such as fine-tuning methods. Future work should extend evaluation across languages, improve contextual stigma detection using more advanced approaches such as LLM-based methods, and determine the clinical significance and downstream effects of stigmatizing language in real-world care.

In conclusion, this study systematically evaluated stigmatizing language in LLM-generated reasoning traces across a large number of real-world clinical tasks. Stigmatizing language was observed across the evaluated models and could be inherited from, generated independently of, or amplified relative to source clinical text. Neither increasing model scale nor medical specialization consistently corresponded to lower stigma, whereas destigmatizing prompt can largely reduce stigma in several examined models but showed model- and task-specific effects. These findings establish stigmatizing language as a measurable and potentially modifiable dimension of clinical AI safety and highlight the need for direct evaluation and robust destigmatization strategies before clinical deployment.

## Methods

### Study Design

Our data source originated from BRIDGE, a comprehensive multilingual benchmark comprising 87 tasks sourced from real-world clinical data sources across 9 languages^34^. For this study, we focused on the subset of 35 tasks in English. The tasks consist of four different characteristics: 3 NLP task types, 15 clinical applications, 7 source clinical document types, and 11 medical applications (**Figure 6**). The 107 models can be categorized into three groups: (a) By training domain: General (n = 81) and medical (n = 26) models; (b) By model licensing: Open-source (n = 102) and proprietary (n = 5) models; (c) By reasoning capability: Reasoning (n = 41) and non-reasoning (n = 66) models (**Figure 6**). **Appendix 1** and **Appendix 3** show the detailed taxonomy of the 35 clinical tasks.

**Figure 6.**
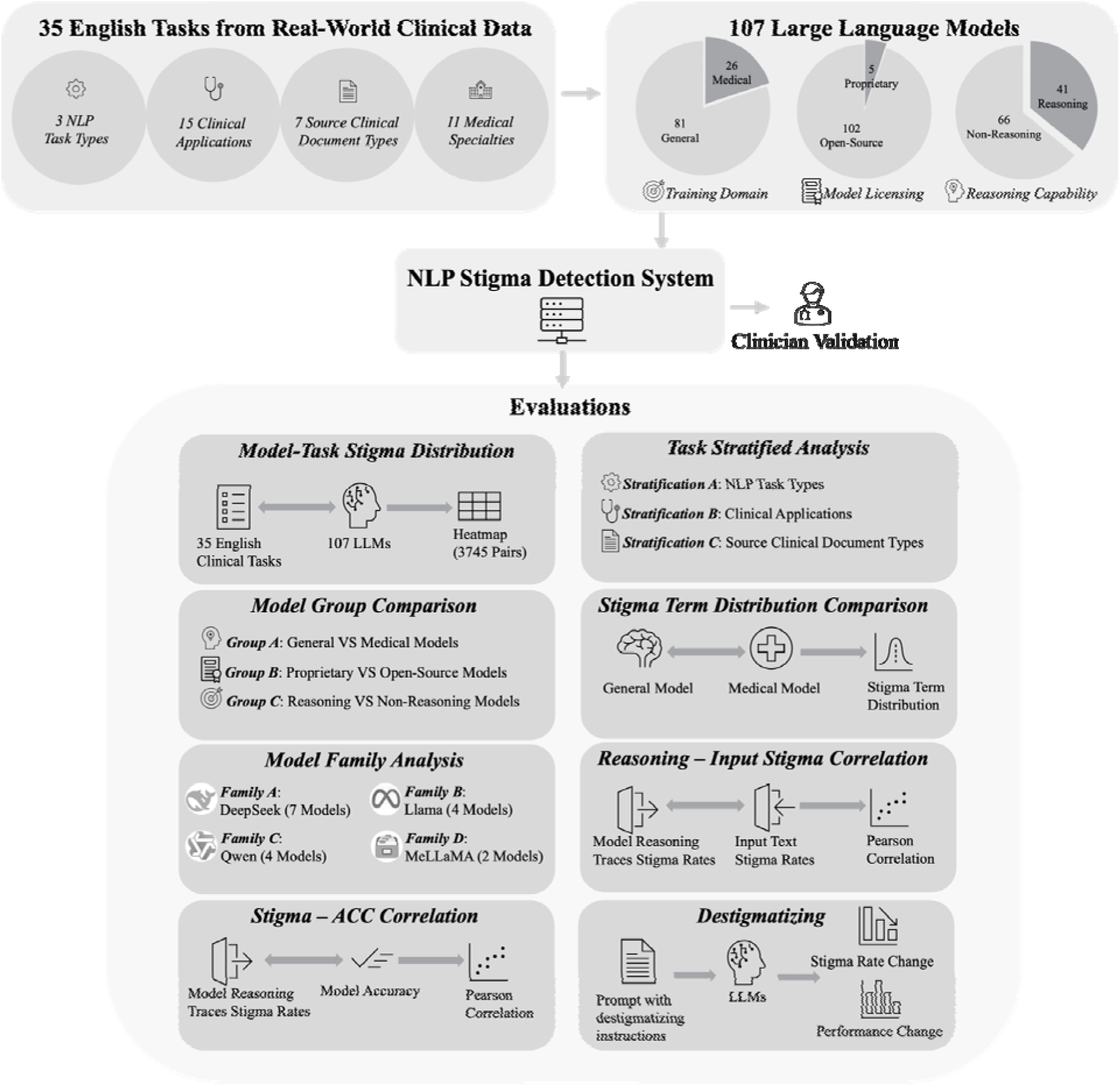
The workflow of this study.

For each task, we evaluated 107 LLMs. Model reasoning traces were obtained from outputs generated under the CoT inference setting, in which each model was prompted to generate an analysis of the question before providing the final answer. **Appendix 4** shows the detailed taxonomy of the 107 models. This yielded a total of 3,745 model-task pairs.

### NLP Stigma Detection System

The NLP stigma detection system has been developed in our previous work, which has been physician-validated to be accurate (NPV 100%, PPV 94.0%) on identifying stigma languages from EHR notes^2–4^. It presets rules that identify stigma terms in sentences without including terms that are associated with special cases. For example, “alcoholic” is a stigma term that will be identified when it is directed towards an individual (e.g., “He is an alcoholic”), but if the word “alcoholic” is associated with the phrase “alcoholic cirrhosis”, a specific illness, then it will not be considered as a stigma term. The system works by splitting each piece of target text into individual sentences and using rule-based matching to detect stigma terms within each sentence, excluding the special cases. Finally, the system assigns a binary label to each sentence, indicating whether it contains stigma terms. For sentences containing stigmatizing language, the system reports the stigma terms detected in each sentence (**Figure 6**). For stigma terms that are substrings of other stigma terms, only the longest term will be detected and counted. After processing, the system outputs a file that records the sentence, the corresponding binary stigma label, the detected stigma terms (if applicable), together with the model-task pair to which the sentence belongs. **Appendix 2** presents the complete list of detected stigma terms, their stigmatizing language categories, and their occurrence counts and proportions. To analyze the correlation between the model’s reasoning traces stigma rate and the corresponding input (i.e., the clinical data) stigma rate, we also applied the stigma detection system on each input sample.

### Validation on the NLP Stigma Detection System

We conducted a structured evaluation of the NLP stigma detection system. Specifically, we randomly sampled 100 positive and 100 negative sentences flagged by the system (Details in **Appendix 6**). Two board-certified psychiatrists (D.B.H. and L.M.) manually reviewed the sentences to determine whether they contained stigmatizing language, using criteria aligned with prior empirical work on stigmatizing language in clinical notes^4^. Discrepant annotations were resolved through discussion to determine the final stigma label. We calculated Cohen’s κ between the two psychiatrists before resolving the discrepant cases, along with the positive predictive value (PPV) and negative predictive value (NPV) of the system’s outputs relative to the annotated cases agreed by the two psychiatrists.

### Stigma Rate Calculation and Distribution Analysis

The stigma identification NLP tool processed the text at a sentence level. For each model–task pair, we defined the stigma rate as:

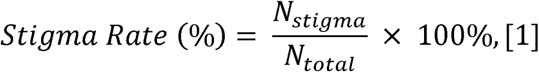

where *N_total_* is the total number of samples in the model-task pair, and *N_stigma_* is the number of samples with at least one stigma term detected in any of its sentences by the stigma detection NLP system.

We computed the reasoning traces and input text stigma rates for each model-task pair using **Equation 1**. We plotted both a heatmap and a histogram showing the overall distribution of the reasoning traces stigma rates for all model-task pairs (**Figure 1**, **Figure 2A**). We then examined the average stigma rate across all clinical applications (**Figure 2B**), NLP task types (**Figure 2C**), and source clinical document types (**Figure 2D**), where the average stigma rate is derived by taking the unweighted mean of the reasoning traces stigma rate of all tasks that belong to this category. If a task has multiple labels for one characteristic, we counted it in each relevant label. For example, tasks with source document type “Discharge Summary, Progress Note” contribute to the stigma rate of both “Discharge Summary” and “Progress Note”.

For the stigma rate analysis across NLP task types and source clinical document types, we selected two best-performing proprietary models (gpt-4o-0806^30^ and gemini-2.5-flash^31^) and two best-performing open-source models (DeepSeek-R1^32^ and gemma-4-31B-it^33^) based on the BRIDGE evaluation results^34^ and compared their stigma rates with the average value of all 107 models.

### Model Group Comparisons

We also compared the average reasoning traces stigma rate within each of the three model groups by taking the unweighted mean of all models’ reasoning traces stigma rates that fall into the respective model type. We performed a two-sided Welch t-test^47^ with unequal variances to determine whether the difference in stigma rates within each model group is statistically significant (**Figure 3A**). To investigate whether the stigma term distribution will vary among the model groups, we checked the distribution differences between general and medical models (**Figure 3B**).

### Relation of Stigma Rate with LLM Model Scales

To assess whether enlarging model scales can help reduce the generation of stigma terms, we selected four different model families (DeepSeek^32^, Llama^39^, Qwen^48^, and MeLLaMA^49^) and tracked the relationship between their parameter sizes and reasoning traces stigma rates (**Figure 4A**). Here, the reasoning traces stigma rate of each model is the unweighted mean of the reasoning traces stigma rate of all 35 tasks on that model.

### Associations Between Reasoning-Trace Stigma, Input-Text Stigma, and Model Performance

We examined two associations involving the stigma rate in model reasoning traces. The first one is the stigma rate between the model’s reasoning traces and the corresponding input text (**Figure 4B**). The second correlation is between the reasoning traces stigma rate and the model’s performance on the tasks (**Figure 4C**). We calculated the Pearson correlation in both cases, together with the statistical significance of the correlation. For the first correlation, we used all 3,745 model-task pairs. For the second correlation, we focused on 15 of the 35 tasks that are classification tasks, as it is more straightforward to quantify the model performance on classification tasks via accuracy. This resulted in 1,605 model-task pairs being involved in the analysis. To show the correlation between model stigma rates and model accuracy, we fit the data with a simple ordinary least squares (OLS) linear regression and calculated the 95% confidence interval band for the mean regression line using the Student’s t-distribution.

### Destigmatizing Experiments

As an experimental approach, we applied prompt engineering to see if it helps reduce model stigma rates. Specifically, we selected one best-performing model from five model families (Mistral^38^, Gemma^33^, Llama^39^, GPT^30^, and Qwen^40^) and reran the CoT inference using an engineering prompt (**Appendix 5**) that deliberately instructs the models to avoid generating stigma terms in their outputs based on the stigma terms list in **Appendix 2**. Aside from comparing the stigma rate differences (**Figure 5A**), we also compared the performance differences of the models before and after applying the new destigmatizing prompt (**Figure 5B**). When comparing the model performance difference by destigmatizing, we only focused on classification (N = 15) and extraction (N = 15) tasks and excluded the generation (N = 5) tasks (see **Appendix 3** “Task Type” column for task types). Because the evaluation of text generation task is multidimensional and task-dependent and there is no single automatic metric is generally accepted as a complete, task-independent measure.^50,51^ Accuracy and the event-level F1 (calculated by micro-scoring, measuring extraction precision and recall across entities and their attributes) are used as the evaluation metrics for the classification and extraction tasks, respectively.

### Ethics Approval and Consent to Participate

This study used de-identified clinical text and model-generated outputs from the BRIDGE benchmark^34^. Because no human participants were involved, ethics approval and informed consent were not required.

### Use of generative AI

The authors acknowledge the use of ChatGPT to assist with grammar checking and language editing of the manuscript. No AI tool was used in the study design, data collection, data analysis, interpretation of results, or generation of scientific conclusions.

## Author contribution

J.Y. designed and supervised this study. Y.Y. and B.G. conducted the experiments and analyzed the results. J.Y., B.G., and Y.Y. drafted the initial manuscript. D.B.H. and L.M. manually reviewed the stigma samples to evaluate the performance of NLP system for stigma detection. J.W. provided support on the data generation of LLM reasoning outputs. L.Z. provided support on the development of the NLP system for stigma detection. R.W., J.B.G, S.L., Q.C., N.L, P.S.W, L.A.C, D.W.B, K.J.L, L.Z and J.Y contributed to manuscript design and refinement. All authors revised, read, and approved the manuscript.

## Acknowledgements

This study was partially funded by PCORI ME-2022C1-25646, Goldberg Scholarship and Brigham Research Institute. The funders had no role in study design, data collection and analysis, decision to publish or preparation of the manuscript.

## Data availability

This study uses clinical text tasks sourced from the BRIDGE benchmark, the detailed dataset information is listed in **Appendix 3**. We release the prompts, together with the models’ outputs and LLM reasoning traces of the tasks that are allowed to be open-sourced. The data can be found at https://github.com/YLab-Open/LLM-Stigma-Language. Additional processed outputs are available from the corresponding authors on reasonable request, where permitted by source-data licenses and privacy restrictions.

## Code availability

The corresponding analysis evaluation code can be found at https://github.com/YLab-Open/LLM-Stigma-Language

