## Supporting Material for "Large Language Models Generate Stigmatizing Language During Reasoning Over Real-World Clinical Data"

**Supplementary**

### Appendix 1: Task taxonomy of the 35 English clinical tasks

Below is the task taxonomy of the 35 English clinical tasks. Distribution of tasks by (A) NLP task type, (B) source clinical document type, (C) clinical specialty, and (D) clinical application. Tasks with multiple labels in a category are counted in each relevant label, so the total counts in some subplots will exceed 35. For detailed definitions of each category under “NLP Task Type”, “Sourced Clinical Document Type”, “Clinical Specialty”, and “Clinical Application”, please refer to BRIDGE^1^.

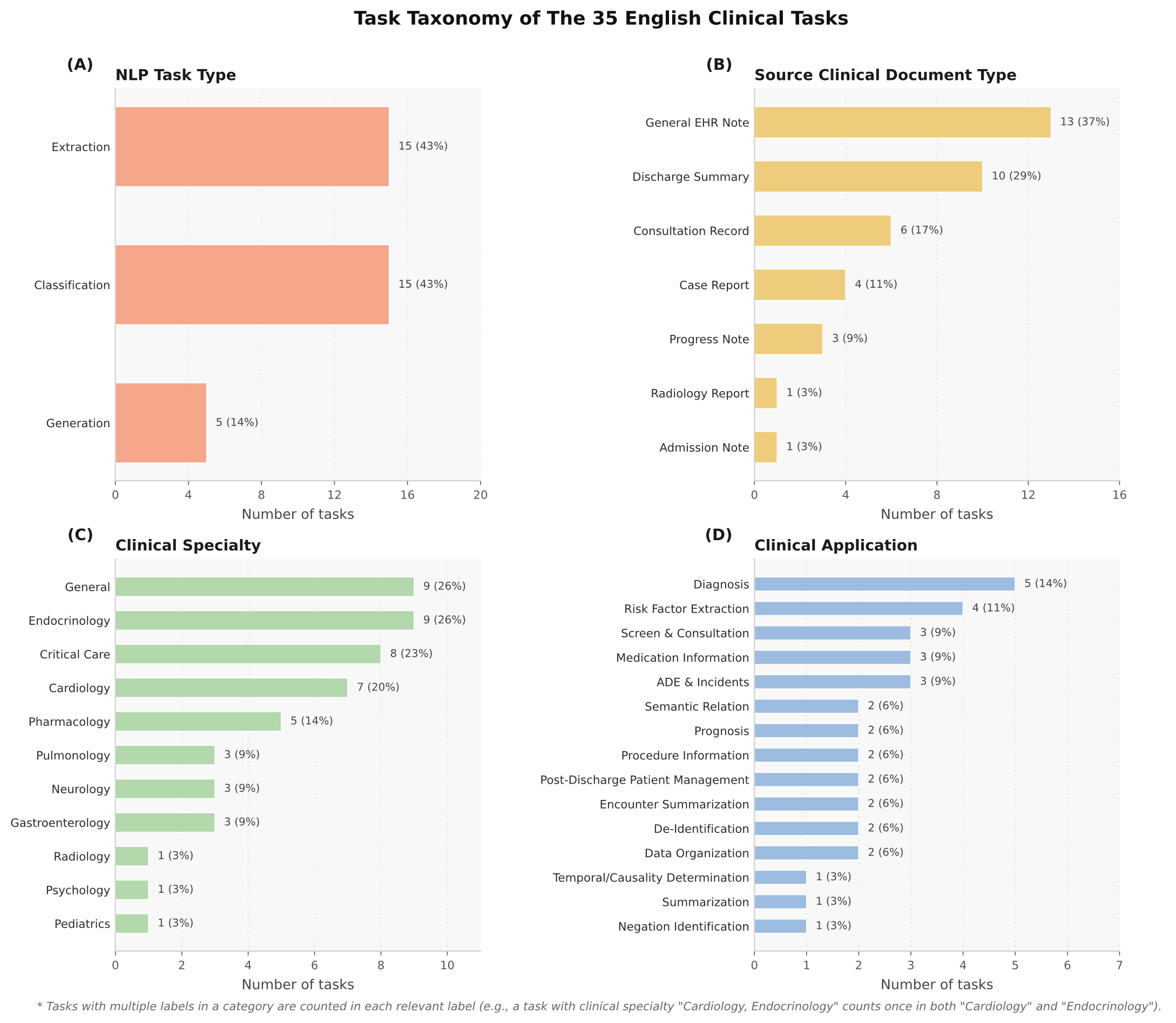

### Appendix 2: Most frequent stigma terms in LLM outputs

Below are the most frequent stigma terms in LLM outputs, sorted by stigma term counts. Terms are ranked by frequency across all model outputs (107 models × 35 tasks; total stigma terms detected =132,706). The "Proportion (%)" column: proportion of the stigma terms over total stigma terms detected. The top three terms ("abuse", "alcoholic”, “drug abuse") account for 90.61% of the detected terms; the top ten terms account for 97.67% of the detected terms.

| **Rank** | **Stigma Term** | **Category** | **Count** | **Proportion (%)** |
| --- | --- | --- | --- | --- |
| 1 | abuse | Substance use / addiction-related | 105,884 | 79.79 |
| 2 | alcoholic | Substance use / addiction-related | 7,314 | 5.51 |
| 3 | drug abuse | Substance use / addiction-related | 7,045 | 5.31 |
| 4 | clean | Descriptor (context-dependent) | 2,449 | 1.85 |
| 5 | replacement therapy | Treatment / regimen terminology | 1,778 | 1.34 |
| 6 | user | Descriptor (context-dependent) | 1,457 | 1.10 |
| 7 | alcoholics | Substance use / addiction-related | 1,393 | 1.05 |
| 8 | addicted | Substance use / addiction-related | 889 | 0.67 |
| 9 | abusers | Substance use / addiction-related | 761 | 0.57 |
| 10 | abusing | Substance use / addiction-related | 644 | 0.49 |
| 11 | abuses | Substance use / addiction-related | 525 | 0.40 |
| 12 | addict | Substance use / addiction-related | 517 | 0.39 |
| 13 | dirty | Descriptor (context-dependent) | 413 | 0.31 |
| 14 | abuser | Substance use / addiction-related | 315 | 0.24 |
| 15 | polysubstance dependence | Substance use / addiction-related | 295 | 0.22 |
| 16 | medication - assisted treatment | Treatment / regimen terminology | 287 | 0.22 |
| 17 | addicts | Substance use / addiction-related | 223 | 0.17 |
| 18 | users | Descriptor (context-dependent) | 194 | 0.15 |
| 19 | substance dependence | Substance use / addiction-related | 110 | 0.08 |
| 20 | drug abuser | Substance use / addiction-related | 87 | 0.07 |
| 21 | opioid substitution | Treatment / regimen terminology | 52 | 0.04 |
| 22 | opioid replacement therapy | Treatment / regimen terminology | 25 | 0.02 |
| 23 | substance abuser | Substance use / addiction-related | 16 | 0.01 |
| 24 | drug abusers | Substance use / addiction-related | 8 | 0.01 |
| 25 | cleans | Descriptor (context-dependent) | 7 | 0.01 |
| 26 | drunk | Descriptor (context-dependent) | 7 | 0.01 |
| 27 | substance abusers | Substance use / addiction-related | 5 | 0.00 |
| 28 | drug abuses | Substance use / addiction-related | 3 | 0.00 |
| 29 | medication assisted treatment | Treatment / regimen terminology | 1 | 0.00 |
| 30 | former addict | Substance use / addiction-related | 1 | 0.00 |
| 31 | junkie | Pejorative label | 1 | 0.00 |

### Appendix 3: Characteristics of the 35 English clinical tasks

Below are the characteristics of the 35 English clinical tasks, sorted alphabetically by “Task Name”. The “Input Text Stigma Rate (%)” column: unweighted mean of the stigma rate on all 107 model input text of the task. The “Reasoning Traces Stigma Rate (%)” column: unweighted mean of the stigma rate on all 107 model output text of the task. See Equation (1) of the main text for the definition of the stigma rate. For detailed definitions of each category under “Task Type”, “Sourced Clinical Document”, “Clinical Specialty”, and “Clinical Application”, please refer to BRIDGE^1^.

| **Task Name** | **Task Type** | **Sourced Clinical Document** | **Clinical Specialty** | **Clinical Application** | **Input Text  Stigma Rate (%)** | **Reasoning Traces Stigma Rate (%)** |
| --- | --- | --- | --- | --- | --- | --- |
| ADE-Drug Dosage | Extraction | Case Report | Pharmacology | Medication information | 1.04 | 0.87 |
| ADE-Extraction | Extraction | Case Report | Pharmacology | ADE & Incidents | 0.47 | 0.54 |
| ADE-Identification | Classification | Case Report | Pharmacology | ADE & Incidents | 0.38 | 0.44 |
| BrainMRI-AIS | Classification | Radiology Report | Neurology, Radiology | Diagnosis | 0.00 | 0.04 |
| ClinicalNotes-UPMC | Classification | General EHR Note | General | Negation identification | 0.00 | 0.01 |
| CLIP | Classification | Discharge Summary | Critical Care | Post-discharge patient management | 0.11 | 0.20 |
| GOUT-CC-Consensus | Classification | Admission Note | Endocrinology | Diagnosis | 0.24 | 0.37 |
| HealthCareMagic-100k | Generation | Consultation Record | General | Screen & Consultation | 0.45 | 0.89 |
| icliniq-10k | Generation | Consultation Record | General | Screen & Consultation | 0.41 | 0.58 |
| Medication Extraction | Extraction | Discharge Summary | Pharmacology | Medication information | 6.11 | 0.31 |
| MEDIQA 2019-RQE | Classification | Consultation Record | General | Screen & Consultation | 0.87 | 0.99 |
| MEDIQA 2023-chat-A | Generation | Consultation Record | General | Encounter summarization | 2.00 | 2.47 |
| MEDIQA 2023-sum-A | Classification | Consultation Record | General | Data organization | 1.01 | 1.08 |
| MEDIQA 2023-sum-B | Generation | Consultation Record | General | Encounter summarization | 1.01 | 1.82 |
| MedNLI | Classification | General EHR Note | Critical Care | Semantic relation | 0.49 | 0.72 |
| MedSTS | Classification | General EHR Note | General | Semantic relation | 0.14 | 0.33 |
| MIMIC-III Outcome.LoS | Classification | Discharge Summary | Critical Care | Prognosis | 14.60 | 8.87 |
| MIMIC-III Outcome.Mortality | Classification | Discharge Summary | Critical Care | Prognosis | 13.30 | 10.23 |
| MIMIC-IV BHC | Generation | General EHR Note | Critical Care | Summarization | 7.90 | 5.07 |
| MIMIC-IV CDM | Classification | General EHR Note | Gastroenterology | Diagnosis | 3.33 | 2.60 |
| MIMIC-IV DiReCT.Dis | Classification | General EHR Note | Cardiology, Gastroenterology, Neurology, Pulmonology, Endocrinology | Diagnosis | 5.98 | 4.30 |
| MIMIC-IV DiReCT.PDD | Classification | General EHR Note | Cardiology, Gastroenterology, Neurology, Pulmonology, Endocrinology | Diagnosis | 5.77 | 4.20 |
| MTSamples | Classification | Case Report | General | Data organization | 4.26 | 1.81 |
| MTSamples-Temporal | Extraction | Discharge Summary | Pediatrics, Psychology | Temporal/Causality determination | 12.41 | 2.26 |
| n2c2 2006-De-identification | Extraction | Discharge Summary | Pulmonology | De-identification | 5.91 | 1.35 |
| n2c2 2010-Assertion | Extraction | Discharge Summary, Progress Note | Critical Care | Post-discharge patient management | 6.72 | 4.40 |
| n2c2 2010-Concept | Extraction | Discharge Summary, Progress Note | Critical Care | Procudure information | 5.71 | 2.32 |
| n2c2 2010-Relation | Extraction | Discharge Summary, Progress Note | Critical Care | Procudure information | 6.78 | 2.96 |
| n2c2 2014-CAD | Extraction | General EHR Note | Cardiology, Endocrinology | Risk factor extraction | 6.64 | 1.06 |
| n2c2 2014-De-identification | Extraction | General EHR Note | Endocrinology | De-identification | 6.23 | 1.66 |
| n2c2 2014-Diabetes | Extraction | General EHR Note | Cardiology, Endocrinology | Risk factor extraction | 7.69 | 0.46 |
| n2c2 2014-Hyperlipidemia | Extraction | General EHR Note | Cardiology, Endocrinology | Risk factor extraction | 6.02 | 0.46 |
| n2c2 2014-Hypertension | Extraction | General EHR Note | Cardiology, Endocrinology | Risk factor extraction | 7.89 | 0.46 |
| n2c2 2014-Medication | Extraction | General EHR Note | Cardiology, Endocrinology | Medication information | 6.65 | 0.71 |
| n2c2 2018-ADE&Medication | Extraction | Discharge Summary | Pharmacology | ADE & Incidents | 11.88 | 1.42 |

### Appendix 4: Characteristics of the 107 evaluated large language models

Below are the characteristics of the 107 evaluated large language models, sorted alphabetically by “Model Name”. The “Domain” column: Medical = The model is specifically fine-tuned on clinical and /or medical corpus; General = The model is not fine-tuned on clinical and/or medical corpus. The “Reasoning” column: Yes = model has thinking capability and thinking capability is enabled during inference; No = model does not support thinking capability or has thinking capability disabled during inference. The “Reasoning Traces Stigma Rate (%)” column: unweighted mean of the stigma rate on the model output text across all 35 tasks. See Equation (1) of the main text for the definition of the stigma rate.

| **Model Name** | **Domain** | **Reasoning** | **License** | **Reasoning Traces Stigma Rate (%)** | **Model URL** |
| --- | --- | --- | --- | --- | --- |
| AntAngelMed | Medical | Yes | Apache 2.0 | 3.30 | https://huggingface.co/MedAIBase/AntAngelMed |
| Athene-V2-Chat | General | No | Nexusflow Research License | 1.59 | https://huggingface.co/Nexusflow/Athene-V2-Chat |
| Baichuan-M1-14B-Instruct | Medical | No | Baichuan-M1-14B | 1.41 | https://huggingface.co/baichuan-inc/Baichuan-M1-14B-Instruct |
| Baichuan-M2-32B | Medical | Yes | Apache 2.0 | 4.24 | https://huggingface.co/baichuan-inc/Baichuan-M2-32B |
| BioMistral-7B | Medical | No | Apache 2.0 | 0.77 | https://huggingface.co/BioMistral/BioMistral-7B |
| Biomni-R0-32B-Preview-Non-Thinking | Medical | Yes | MIT | 1.65 | https://huggingface.co/biomni/Biomni-R0-32B-Preview |
| Biomni-R0-32B-Preview-Thinking | Medical | Yes | MIT | 2.55 | https://huggingface.co/biomni/Biomni-R0-32B-Preview |
| DeepSeek-R1 | General | Yes | MIT | 2.17 | https://huggingface.co/deepseek-ai/DeepSeek-R1 |
| DeepSeek-R1-0528-Qwen3-8B | General | Yes | MIT | 3.42 | https://huggingface.co/deepseek-ai/DeepSeek-R1-0528-Qwen3-8B |
| DeepSeek-R1-Distill-Llama-70B | General | Yes | MIT | 2.11 | https://huggingface.co/deepseek-ai/DeepSeek-R1-Distill-Llama-70B |
| DeepSeek-R1-Distill-Llama-8B | General | Yes | MIT | 1.90 | https://huggingface.co/deepseek-ai/DeepSeek-R1-Distill-Llama-8B |
| DeepSeek-R1-Distill-Qwen-1.5B | General | Yes | MIT | 2.06 | https://huggingface.co/deepseek-ai/DeepSeek-R1-Distill-Qwen-1.5B |
| DeepSeek-R1-Distill-Qwen-14B | General | Yes | MIT | 1.88 | https://huggingface.co/deepseek-ai/DeepSeek-R1-Distill-Qwen-14B |
| DeepSeek-R1-Distill-Qwen-32B | General | Yes | MIT | 2.09 | https://huggingface.co/deepseek-ai/DeepSeek-R1-Distill-Qwen-32B |
| DeepSeek-R1-Distill-Qwen-7B | General | Yes | MIT | 2.40 | https://huggingface.co/deepseek-ai/DeepSeek-R1-Distill-Qwen-7B |
| gemini-1.5-pro | General | No | Proprietary | 1.54 | https://deepmind.google/technologies/gemini/ |
| gemini-2.0-flash | General | No | Proprietary | 1.38 | https://deepmind.google/technologies/gemini/ |
| gemini-2.5-flash | General | Yes | Proprietary | 1.64 | https://deepmind.google/technologies/gemini/ |
| gemma-2-27b-it | General | No | Gemma | 1.43 | https://huggingface.co/google/gemma-2-27b-it |
| gemma-2-9b-it | General | No | Gemma | 1.50 | https://huggingface.co/google/gemma-2-9b-it |
| gemma-3-12b-it | General | No | Gemma | 1.59 | https://huggingface.co/google/gemma-3-12b-it |
| gemma-3-1b-it | General | No | Gemma | 2.11 | https://huggingface.co/google/gemma-3-1b-it |
| gemma-3-27b-it | General | No | Gemma | 1.55 | https://huggingface.co/google/gemma-3-27b-it |
| gemma-3-4b-it | General | No | Gemma | 1.92 | https://huggingface.co/google/gemma-3-4b-it |
| gemma-4-26B-A4B-it | General | No | Apache 2.0 | 1.10 | https://huggingface.co/google/gemma-4-26B-A4B-it |
| gemma-4-31B-it | General | No | Apache 2.0 | 1.36 | https://huggingface.co/google/gemma-4-31B-it |
| gemma-4-E2B-it | General | No | Apache 2.0 | 1.32 | https://huggingface.co/google/gemma-4-E2B-it |
| gemma-4-E4B-it | General | No | Apache 2.0 | 1.28 | https://huggingface.co/google/gemma-4-E4B-it |
| gpt-35-turbo | General | No | Proprietary | 1.79 | https://platform.openai.com/docs/models/gpt-3.5-turbo |
| gpt-4o | General | No | Proprietary | 1.67 | https://platform.openai.com/docs/models/gpt-4o |
| gpt-oss-120b | General | Yes | Apache 2.0 | 2.63 | https://huggingface.co/openai/gpt-oss-120b |
| gpt-oss-20b | General | Yes | Apache 2.0 | 2.68 | https://huggingface.co/openai/gpt-oss-20b |
| HuatuoGPT-o1-70B | Medical | Yes | Apache 2.0 | 1.22 | https://huggingface.co/FreedomIntelligence/HuatuoGPT-o1-70B |
| HuatuoGPT-o1-72B | Medical | Yes | Apache 2.0 | 1.29 | https://huggingface.co/FreedomIntelligence/HuatuoGPT-o1-72B |
| HuatuoGPT-o1-7B | Medical | Yes | Apache 2.0 | 1.48 | https://huggingface.co/FreedomIntelligence/HuatuoGPT-o1-7B |
| HuatuoGPT-o1-8B | Medical | Yes | Apache 2.0 | 1.51 | https://huggingface.co/FreedomIntelligence/HuatuoGPT-o1-8B |
| Hulu-Med-14B | Medical | Yes | Apache 2.0 | 1.47 | https://huggingface.co/ZJU-AI4H/Hulu-Med-14B |
| Hulu-Med-32B | Medical | Yes | Apache 2.0 | 1.64 | https://huggingface.co/ZJU-AI4H/Hulu-Med-32B |
| Hulu-Med-7B | Medical | Yes | Apache 2.0 | 1.76 | https://huggingface.co/ZJU-AI4H/Hulu-Med-7B |
| K2-Think | General | Yes | Apache 2.0 | 2.42 | https://huggingface.co/LLM360/K2-Think |
| Llama-2-13b-chat | General | No | llama2 | 1.73 | https://huggingface.co/meta-llama/Llama-2-13b-chat-hf |
| Llama-2-70b-chat | General | No | llama2 | 2.16 | https://huggingface.co/meta-llama/Llama-2-70b-chat-hf |
| Llama-2-7b-chat | General | No | llama2 | 1.69 | https://huggingface.co/meta-llama/Llama-2-7b-chat-hf |
| Llama-3-70B-UltraMedical | Medical | No | Llama-3 | 2.07 | https://huggingface.co/TsinghuaC3I/Llama-3-70B-UltraMedical |
| Llama-3.1-70B-Instruct | General | No | Llama-3.1 | 1.54 | https://huggingface.co/meta-llama/Llama-3.1-70B-Instruct |
| Llama-3.1-8B-Instruct | General | No | Llama-3.1 | 1.80 | https://huggingface.co/meta-llama/Llama-3.1-8B-Instruct |
| Llama-3.1-8B-UltraMedical | Medical | No | Llama-3 | 3.03 | https://huggingface.co/TsinghuaC3I/Llama-3.1-8B-UltraMedical |
| Llama-3.1-Nemotron-70B-Instruct-HF | General | No | Llama-3.1 | 1.52 | https://huggingface.co/nvidia/Llama-3.1-Nemotron-70B-Instruct-HF |
| Llama-3.2-1B-Instruct | General | No | Llama-3.1 | 2.03 | https://huggingface.co/meta-llama/Llama-3.2-1B-Instruct |
| Llama-3.2-3B-Instruct | General | No | Llama-3.1 | 1.74 | https://huggingface.co/meta-llama/Llama-3.2-3B-Instruct |
| Llama-3.3-70B-Instruct | General | No | Llama-3.3 | 1.81 | https://huggingface.co/meta-llama/Llama-3.3-70B-Instruct |
| Llama-4-Scout-17B-16E-Instruct | General | No | Llama-4 | 1.93 | https://huggingface.co/meta-llama/Llama-4-Scout-17B-16E-Instruct |
| Llama3-OpenBioLLM-70B | Medical | No | Llama-3 | 1.40 | https://huggingface.co/aaditya/Llama3-OpenBioLLM-70B |
| Llama3-OpenBioLLM-8B | Medical | No | Llama-3 | 1.13 | https://huggingface.co/aaditya/Llama3-OpenBioLLM-8B |
| Magistral-Small-2506 | General | Yes | Apache 2.0 | 1.68 | https://huggingface.co/mistralai/Magistral-Small-2506 |
| medgemma-27b-it | Medical | No | Health AI Developer Foundations terms of use | 2.38 | https://huggingface.co/google/medgemma-27b-it |
| medgemma-27b-text-it | Medical | No | Health AI Developer Foundations terms of use | 2.37 | https://huggingface.co/google/medgemma-27b-text-it |
| medgemma-4b-it | Medical | No | Health AI Developer Foundations terms of use | 1.75 | https://huggingface.co/google/medgemma-4b-it |
| meditron-70b | Medical | No | Apache 2.0 | 2.15 | https://github.com/epfLLM/meditron |
| meditron-7b | Medical | No | Apache 2.0 | 1.72 | https://github.com/epfLLM/meditron |
| MedReason-8B | Medical | Yes | Apache 2.0 | 1.54 | https://huggingface.co/UCSC-VLAA/MedReason-8B |
| MeLLaMA-13B-chat | Medical | No | PhysioNet Credentialed Health Data License 1.5.0 | 0.43 | https://physionet.org/content/me-llama/1.0.0/ |
| MeLLaMA-70B-chat | Medical | No | PhysioNet Credentialed Health Data License 1.5.0 | 0.45 | https://physionet.org/content/me-llama/1.0.0/ |
| Ministral-8B-Instruct-2410 | General | No | MRL | 1.67 | https://huggingface.co/mistralai/Ministral-8B-Instruct-2410 |
| Mistral-Large-Instruct-2411 | General | No | MRL | 1.70 | https://huggingface.co/mistralai/Mistral-Large-Instruct-2411 |
| Mistral-Small-24B-Instruct-2501 | General | No | Apache 2.0 | 1.25 | https://huggingface.co/mistralai/Mistral-Small-24B-Instruct-2501 |
| Mistral-Small-3.1-24B-Instruct-2503 | General | No | Apache 2.0 | 1.64 | https://huggingface.co/mistralai/Mistral-Small-3.1-24B-Instruct-2503 |
| Mistral-Small-Instruct-2409 | General | No | MRL | 1.45 | https://huggingface.co/mistralai/Mistral-Small-Instruct-2409 |
| MMed-Llama-3-8B | Medical | No | Llama-3 | 1.99 | https://huggingface.co/Henrychur/MMed-Llama-3-8B |
| OpenThinker3-7B | General | Yes | Apache 2.0 | 2.72 | https://huggingface.co/open-thoughts/OpenThinker3-7B |
| Phi-3.5-mini-instruct | General | No | MIT | 1.65 | https://huggingface.co/microsoft/Phi-3.5-mini-instruct |
| Phi-3.5-MoE-instruct | General | No | MIT | 1.59 | https://huggingface.co/microsoft/Phi-3.5-MoE-instruct |
| Phi-4 | General | No | MIT | 1.77 | https://huggingface.co/microsoft/phi-4 |
| Phi-4-mini-instruct | General | No | MIT | 1.20 | https://huggingface.co/microsoft/Phi-4-mini-instruct |
| Phi-4-mini-reasoning | General | Yes | MIT | 2.76 | https://huggingface.co/microsoft/Phi-4-mini-reasoning |
| Qwen2.5-1.5B-Instruct | General | No | Apache 2.0 | 1.62 | https://huggingface.co/Qwen/Qwen2.5-1.5B-Instruct |
| Qwen2.5-32B-Instruct | General | No | Apache 2.0 | 1.83 | https://huggingface.co/Qwen/Qwen2.5-32B-Instruct |
| Qwen2.5-3B-Instruct | General | No | Apache 2.0 | 1.42 | https://huggingface.co/Qwen/Qwen2.5-3B-Instruct |
| Qwen2.5-72B-Instruct | General | No | Qwen | 1.94 | https://huggingface.co/Qwen/Qwen2.5-72B-Instruct |
| Qwen2.5-7B-Instruct | General | No | Apache 2.0 | 1.68 | https://huggingface.co/Qwen/Qwen2.5-7B-Instruct |
| Qwen3-0.6B-Non-Thinking | General | No | Apache 2.0 | 1.58 | https://huggingface.co/Qwen/Qwen3-0.6B |
| Qwen3-0.6B-Thinking | General | Yes | Apache 2.0 | 1.76 | https://huggingface.co/Qwen/Qwen3-0.6B |
| Qwen3-1.7B-Non-Thinking | General | No | Apache 2.0 | 1.98 | https://huggingface.co/Qwen/Qwen3-1.7B |
| Qwen3-1.7B-Thinking | General | Yes | Apache 2.0 | 2.37 | https://huggingface.co/Qwen/Qwen3-1.7B |
| Qwen3-14B-Non-Thinking | General | No | Apache 2.0 | 2.08 | https://huggingface.co/Qwen/Qwen3-14B |
| Qwen3-14B-Thinking | General | Yes | Apache 2.0 | 2.43 | https://huggingface.co/Qwen/Qwen3-14B |
| Qwen3-235B-A22B-Non-Thinking | General | No | Apache 2.0 | 1.88 | https://huggingface.co/Qwen/Qwen3-235B-A22B |
| Qwen3-235B-A22B-Thinking | General | Yes | Apache 2.0 | 3.14 | https://huggingface.co/Qwen/Qwen3-235B-A22B |
| Qwen3-30B-A3B-Instruct-2507 | General | No | Apache 2.0 | 2.33 | https://huggingface.co/Qwen/Qwen3-30B-A3B-Instruct-2507 |
| Qwen3-30B-A3B-Non-Thinking | General | No | Apache 2.0 | 2.01 | https://huggingface.co/Qwen/Qwen3-30B-A3B |
| Qwen3-30B-A3B-Thinking | General | Yes | Apache 2.0 | 2.36 | https://huggingface.co/Qwen/Qwen3-30B-A3B |
| Qwen3-30B-A3B-Thinking-2507 | General | Yes | Apache 2.0 | 2.68 | https://huggingface.co/Qwen/Qwen3-30B-A3B-Thinking-2507 |
| Qwen3-32B-Non-Thinking | General | No | Apache 2.0 | 1.89 | https://huggingface.co/Qwen/Qwen3-32B |
| Qwen3-32B-Thinking | General | Yes | Apache 2.0 | 2.35 | https://huggingface.co/Qwen/Qwen3-32B |
| Qwen3-4B-Instruct-2507 | General | No | Apache 2.0 | 2.26 | https://huggingface.co/Qwen/Qwen3-4B-Instruct-2507 |
| Qwen3-4B-Non-Thinking | General | No | Apache 2.0 | 1.75 | https://huggingface.co/Qwen/Qwen3-4B |
| Qwen3-4B-Thinking | General | Yes | Apache 2.0 | 2.25 | https://huggingface.co/Qwen/Qwen3-4B |
| Qwen3-4B-Thinking-2507 | General | Yes | Apache 2.0 | 3.67 | https://huggingface.co/Qwen/Qwen3-4B-Thinking-2507 |
| Qwen3-8B-Non-Thinking | General | No | Apache 2.0 | 2.13 | https://huggingface.co/Qwen/Qwen3-8B |
| Qwen3-8B-Thinking | General | Yes | Apache 2.0 | 2.51 | https://huggingface.co/Qwen/Qwen3-8B |
| Qwen3-Next-80B-A3B-Instruct | General | No | Apache 2.0 | 2.31 | https://huggingface.co/Qwen/Qwen3-Next-80B-A3B-Instruct |
| Qwen3-Next-80B-A3B-Thinking | General | Yes | Apache 2.0 | 4.23 | https://huggingface.co/Qwen/Qwen3-Next-80B-A3B-Thinking |
| QwenLong-L1-32B | General | Yes | Apache 2.0 | 2.10 | https://huggingface.co/Tongyi-Zhiwen/QwenLong-L1-32B |
| QWQ-32B | General | Yes | Apache 2.0 | 2.42 | https://huggingface.co/Qwen/QwQ-32B |
| QwQ-32B-Preview | General | Yes | Apache 2.0 | 3.83 | https://huggingface.co/Qwen/QwQ-32B-Preview |
| Yi-1.5-34B-Chat-16K | General | No | Apache 2.0 | 1.93 | https://huggingface.co/01-ai/Yi-1.5-34B-Chat-16K |
| Yi-1.5-9B-Chat-16K | General | No | Apache 2.0 | 1.67 | https://huggingface.co/01-ai/Yi-1.5-9B-Chat-16K |

### Appendix 5: Prompt used for destigmatizing

Below is the prompt used for destigmatizing. The list of stigma terms is based on **Appendix 2**. Please note that this part is inserted into the original system prompt fed to the model, so the model’s output format will not change.

*Please do not generate or use any of the following stigma terms anywhere in your response, even in your references to the input text or your internal thinking process:*

*abuse*

*alcoholic*

*drug abuse*

*clean*

*replacement therapy*

*user*

*alcoholics*

*addicted*

*abusers*

*abusing*

*abuses*

*addict*

*dirty*

*abuser*

*polysubstance dependence*

*medication - assisted treatment*

*addicts*

*users*

*substance dependence*

*drug abuser*

*opioid substitution*

*opioid replacement therapy*

*substance abuser*

*drug abusers*

*cleans*

*drunk*

*substance abusers*

*drug abuses*

*medication assisted treatment*

*former addict*

*junkie*

The format of the prompt with or without the destigmatizing patch is like the following:

***No destigmatizing:***

*[Task specific instruction]*

*[Output format instruction]*

***Destigmatizing:***

*[Task specific instruction]*

*[Destigmatizing patch (Please do not generate…)]*

*[Output format instruction]*

### Appendix 6: Data selection process for human review

We gathered the output of the NLP stigma detection system of all LLMs on all tasks and randomly selected 100 positive and 100 negative cases based on the model’s prediction without stratifying by LLMs or tasks, so there may be multiple samples from the same LLM or task, and there will be models or tasks that are not sampled. Since the model splits the input text into sentences and predicts at the sentence level, the sampled data consists of 100 sentences that, based on the NLP system’s prediction, contain at least one predefined stigma term, and 100 sentences that, based on the NLP system’s prediction, do not contain any predefined stigma term.
